# Local Exhaust Ventilation to Control Dental Aerosols and Droplets

**DOI:** 10.1101/2021.07.21.21260917

**Authors:** James R Allison, Christopher Dowson, Kimberley Pickering, Greta Červinskytė, Justin Durham, Nicholas S Jakubovics, Richard Holliday

**Author notes:** **Corresponding author:** Mr James Allison, School of Dental Sciences, Faculty of Medical Sciences, Newcastle University, Framlington Place, Newcastle Upon Tyne, NE2 4BW.

## Abstract

Dental procedures produce aerosols which may remain suspended and travel significant distances from the source. Dental aerosols and droplets contain oral microbes and there is potential for infectious disease transmission and major disruption to dental services during infectious disease outbreaks. One method to control hazardous aerosols often used in industry is Local Exhaust Ventilation (LEV). The aim of this study was to investigate the effect of LEV on aerosols and droplets produced during dental procedures. Experiments were conducted on dental mannequins in an 825.4 m^3^ open plan clinic, and a 49.3 m^3^ single surgery. 10-minute crown preparations were performed with an air-turbine handpiece in the open plan clinic, and 10-minute full mouth ultrasonic scaling in the single surgery. Fluorescein was added to instrument irrigation reservoirs as a tracer. In both settings, Optical Particle Counters (OPCs) were used to measure aerosol particles between 0.3 – 10.0 μm and liquid cyclone air samplers were used to capture aerosolised fluorescein tracer. Additionally, in the open plan setting fluorescein tracer was captured by passive settling onto filter papers in the environment. Tracer was quantified fluorometrically. An LEV device with High Efficiency Particulate Air (HEPA) filtration and a flow rate of 5,000 L/min was used. LEV reduced aerosol production from the air-turbine handpiece by 90% within 0.5 m, and this was 99% for the ultrasonic scaler. OPC particle counts were substantially reduced for both procedures, and air-turbine settled droplet detection reduced by 95% within 0.5 m. The effect of LEV was substantially greater than suction alone for the air-turbine and was similar to the effect of suction for the ultrasonic scaler. LEV reduces aerosol and droplet contamination from dental procedures by at least 90% in the breathing zone of the operator and it is therefore a valuable tool to reduce the dispersion of dental aerosols.

## Introduction

Dental procedures produce aerosols and droplets containing oral microbes (Meethil et al. 2021; Zemouri et al. 2020b). This is relevant during infectious disease outbreaks, where concerns over pathogen dissemination (e.g., SARS-CoV-2) my disrupt dental service provision and pose and infection risk to staff and patients. Pathogen dispersion during Aerosol-Generating Procedures (AGPs) is also an issue in wider healthcare, for example during endotracheal intubation (Tran et al. 2012).

The literature relating to airborne disease transmission has been subject to recent scrutiny, and although it is frequently stated droplets >5 μm diameter do not remain airborne (WHO 2014), this has been questioned by some (Tang et al. 2021). In fact, there is evidence that that droplets 60 – 100 μm remain suspended, thus posing an inhalation risk (Xie et al. 2007).

Several methods of reducing risks from aerosol dispersion in dentistry have been proposed, for example reducing aerosol production using alternative dental handpiece designs (Allison et al. 2021b; Vernon et al. 2021), reducing pathogenic load with mouthrinses or antimicrobial irrigants, and reducing escape from the mouth using dental dam (SDCEP 2020). Where potentially contaminated aerosols do escape, ventilation is important in reducing exposure (Zemouri et al. 2020a), however this is often dictated by building configuration and may be difficult or costly to change. An alternative approach is to use filtration to increase the effective air-exchange rate. This can be achieved using free-standing high-efficiency particulate air (HEPA) filtration devices, however the effect of these is likely to depend on distance from the source, and airflow in the room (Ren et al. 2020).

Local Exhaust Ventilation (LEV) systems are an alternative approach (SDCEP 2021) and capture aerosols at the source, reducing escape into the environment. LEV is used in industry to protect workers from exposure to dust, fumes, and gases during tasks including welding and soldering (HSE 2017). These devices have been referred to in the dental literature as “extra-oral suction/scavenging”, however LEV is a more correct term and is used throughout this paper. Previous studies of LEV for dental procedures have reported promising findings, however to our knowledge, none have evaluated both settled droplets and suspended aerosols together (Ehtezazi et al. 2021; Shahdad et al. 2020). Whilst many dental settings constitute enclosed, single operatories, a proportion of care is delivered in large, open plan clinics, often in educational institutions. The ability of LEV to control the dispersion of aerosols across a large clinical area, thereby reducing exposure to individuals at distant sites is unknown. The aim of this study is to investigate the effect of LEV on the distribution of aerosols and droplets produced during dental procedures.

## Materials and Methods

### Setting

#### Open plan setting

Experiments using an air-turbine handpiece were conducted in an 825.4 m^3^ clinical teaching laboratory at the School of Dental Sciences, Newcastle University, UK, with a supply and extract Heating, Ventilation and Air Conditioning (HVAC) system providing 6.5 Air Changes per Hour (ACH; assessed by an external engineering contractor) via ceiling vents. Each air exchange reduces contaminants by around 63%, therefore after 6 air changes 99.7% of airborne contaminants will be removed, assuming emission has ceased (SDCEP 2021). A rig was constructed around a dental mannequin as previously described (Allison et al. 2021a) comprising platforms spaced at 0.5 m intervals along eight, 4 m, rods, at 45° intervals supported by a central hub (figure 1). This created an 8 m diameter circle around the mannequin, with a centre 28 cm superior to the mannequin’s mouth and 73 cm above the floor. All windows and doors remained closed and only the operator and assistant were present inside the experimental area, leaving immediately after completing the procedure. We used the air-turbine handpiece in the open plan setting as our previous data suggest that this instrument produces widespread contamination which may not be represented in a single surgery setting, compared to the ultrasonic scaler (Allison et al. 2021a).

**Figure 1.**
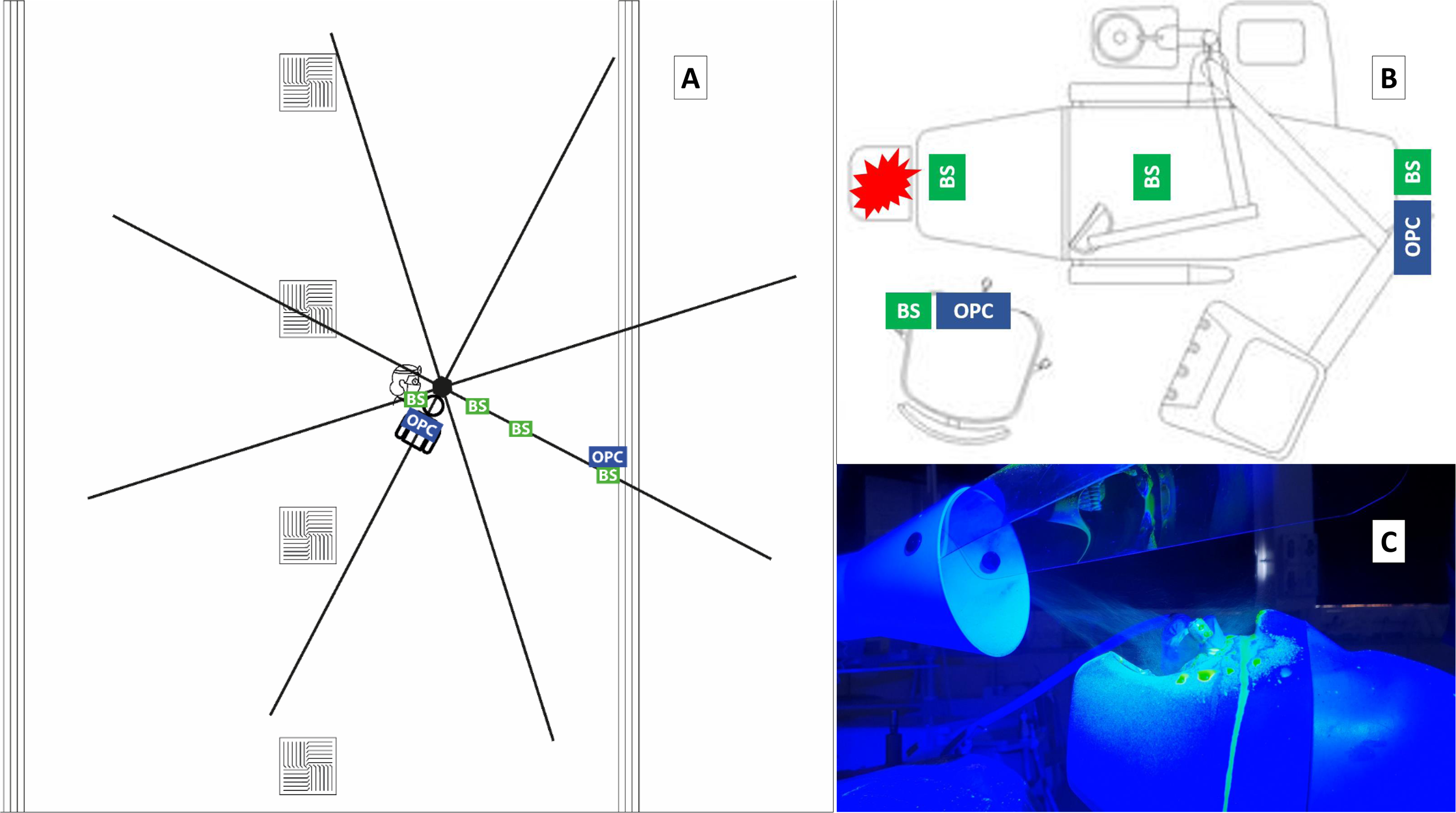
Overview of experimental setup A: Plan view of open plan setting. Sampling locations are shown as boxes (OPC: Optical Particle Counter; BS: BioSampler). The position of air vents in the open plan setting are shown: square vents = air intake; long vents = air output. A rig to support filter papers is shown as black lines radiating from a centre above the mannequin. Filter papers were spaced at 0.5 m intervals on each of the eight rods. B: Plan view of single surgery setting as above. The star indicates the location of the aerosol generating procedure. C: Positioning of the LEV device in relation to the dental mannequin. Further images are presented in the supplementary appendix.

#### Single surgery setting

Experiments using an ultrasonic scaler were conducted in a 49.3 m^3^ enclosed dental surgery at Newcastle Dental Hospital, Newcastle upon Tyne Hospitals NHS Foundation Trust, UK. This setting had a supply and extract HVAC system providing 5.0 ACH via ceiling vents. Only the operator and assistant were present inside the experimental area and left immediately after completing the procedure.

### Pilot testing

Preliminary experiments undertaken to confirm fluorescein is captured by LEV are described in the supplementary appendix.

### Dental procedures

#### Air-turbine handpiece

In the open plan setting, experiments were conducted on a dental simulator unit (Model 4820, A-dec; OR, USA) with a mannequin containing model teeth (Frasaco GmbH; Tettnang, Germany). The mouth of the mannequin was positioned 83 cm above the floor. One operator (RH, height: 170 cm) completed an anterior crown preparation of the upper right central incisor for ten minutes using a air-turbine handpiece (Synea TA-98, W&H (UK) Ltd.; St Albans, UK). The coolant flow rate was 29.3 mL/min. 2.65 mM solution fluorescein sodium tracer was introduced into the irrigation reservoir of the dental unit. In all experiments in this setting, an assistant operated dental suction with an 8.3 mm internal diameter suction tip at a flow rate of 133 L/min of air measured using a flow meter (Ramvac Flowcheck, DentalEZ; PA, USA); this equates to “medium volume suction” according to UK guidelines (NHS Estates 2003). Three replicates were conducted for each experiment as well as for a negative control condition where no procedure was occurring.

#### Ultrasonic scaler

In the single surgery setting, a dental mannequin (P-6/3 TSE, Frasaco GmbH; Tettnang, Germany) was attached to a dental chair (Pelton and Crane Spirit Series, Charlotte, USA) with the mouth positioned 90 cm above the floor. One of two operators (RH, height: 170 cm; GC, height: 169 cm) completed full mouth ultrasonic scaling for ten minutes using a magnetostrictive scaler (Cavitron Select SPS, 30K FSI-1000-94 insert, Dentsply Sirona; PA, USA) at full power (coolant flow rate: 38.6 mL/min). Fluorescein tracer was used as described above. In some experiments, an assistant operated dental suction with a 14.0 mm internal diameter suction tip at a flow rate of 251 L/min of air; this equates to “high volume suction” (NHS Estates 2003). Three replicates were conducted for each experiment as well as for a negative control condition.

### Local Exhaust Ventilation

A DentalAIR UVC AGP Filtration system (DA-UVC1001; VODEX Ltd., UK) was used as the LEV device. This device uses a HEPA filter compliant with EN1822 standards at an air flow rate of 5,000 L/min of air and includes a 254 nm, 27 mW/cm^2^ UVC source in the airflow before filtration. The centre of the device’s inlet nozzle was positioned 10 cm inferior to the chin of the mannequin, and 4 cm above the plane of the mannequin’s mouth (Figure 1 and supplementary appendix)

### Aerosol and droplet detection

#### Optical particle counters

In both settings, two laser-diode optical particle counters (OPCs; 3016 IAQ, Lighthouse; OR, USA) were used to measure suspended aerosols. OPCs had six particle-size channels (0.3, 0.5, 1.0, 2.5, 5.0, and 10.0 μm) with a sampling flow rate of 2.83 L/min and were calibrated by the manufacturer to ISO 21501-4. Instruments sampled continuously at 5-second intervals beginning 2 minutes before the procedure, continuing during the 10-minute procedure, and for 20 minutes after (32 minutes total). OPCs were placed in two positions during each experiment (Figure 1 and supplementary appendix). In the open plan setting, this was 0.5 m inferior to the mouth of the mannequin, and to the left of the mannequin at 2 m; in the single surgery setting, this was 0.5 m to the right of the mannequin, and at 2 m at the foot of the dental chair. Both OPCs were positioned with sampling nozzles 87 cm above the floor. Data were presented as normalised particle counts (particles/ m^3^) over the time-course of the experiment and total particle counts were summed across all particle size channels. As experiments were conducted in real clinical settings, background particle counts were variable. All OPC data were therefore normalised to an internal baseline by subtracting the average counts during the 2 minutes before the procedure from all particle counts. These instruments also measured temperature and relative humidity.

#### Active air sampling

In both settings, liquid cyclone air samplers (BioSampler, SKC Inc.; PA, USA) were placed in four positions during each experiment (Figure 1 and supplementary appendix), after cleaning by alternating washing with distilled water and 70% ethanol to eliminate fluorescein carry-over. In the open plan setting BioSamplers were positioned in the left chest pocket of the operator, and at 0.5, 1.0, and 2.0 m to the left of the mannequin; in the single surgery setting this was at 0.15 m on the mannequin (chest), 0.5 m to the right of the mannequin, and at 1.0 and 2.0 on the dental chair. 20 mL of distilled water was added to the sampling vessels before operation. BioSamplers were operated at an air flow rate of 12.5 L/min using a sampling pump (BioLite+, SKC Inc.; PA, USA) and calibrated using a rotameter (SKC Inc.; PA, USA). Sampling duration was as for OPCs.

100 μL of the sampled solution was then added to wells of a black 96-well microtitre plate with a micro-clear bottom (Greiner Bio-One; NC, USA) in triplicate. Fluorescence was measured using a Synergy HT Microplate Reader (BioTek; VT, USA) at an excitation wavelength of 485 ± 20 nm and emission wavelength of 528 ± 20 nm with the top optical probe. Negative controls were collected and analysed in the same way. The mean (SD) fluorescence reading from negative controls in the open plan setting was 25.2 (1.7) Relative Fluorescence Units (RFU), *n* = 12. In the single surgery setting, this was 25.8 (2.8) RFU, *n* = 12. These values were subtracted from all data for background correction.

#### Passive sampling

This method was used only in the open plan setting for experiments using the air-turbine handpiece. 30 mm diameter grade 1 cotton-cellulose qualitative filter papers (Whatman; Cytiva, MA, USA) were placed onto platforms on the rig surrounding the mannequin prior to each experiment following cleaning of platforms with 70% ethanol. Filter papers were collected following experiments with clean tweezers and placed into individual polypropylene bags. Previous work showed that this eliminates carry-over of fluorescein (Allison et al. 2021a). Fluorescein was recovered by adding 350 μL deionised water. Immersed samples were shaken for 5 minutes at 300 rpm using an orbital shaker at room temperature. Fluorescein was eluted by centrifugation at 15,890 *g* for 3 minutes using a microcentrifuge. 100 μL of the supernatant was transferred to a black 96-well microtitre plate with a micro-clear bottom (Greiner Bio-One; NC, USA) in triplicate to measure fluorescence using the plate reader as for BioSampler samples.

### Statistical methods

Data were collected using Excel (2016, Microsoft; WA, USA) and analysed with SPSS (version24, IBM Corp.; NY, USA) using descriptive statistics.

## Results

Full datasets are available at https://doi.org/10.25405/data.ncl.14987574.

### Open plan setting with air-turbine handpiece

The mean (SD; minimum – maximum) temperature was 23.7 °C (0.5; 22.6 – 25.1 °C) and the relative humidity was 28.8 % (6.3; 20.0 – 38.6 %).

#### Active sampling with optical particle counters

OPC data were collected at two positions in the open plan setting: 0.5 m to sample aerosols in the breathing zone of the operator and assistant, and at 2 m to sample aerosols at the minimum distance between dental chairs recommended by current UK infection prevention and control guidance (Public Health England 2020). Particle counts were substantially lower during all conditions at 2 m compared to at 0.5 m. At both 0.5 m and 2 m, the use of LEV was associated with a substantial reduction in particle counts. Figure 2 shows illustrative data from one repetition at 0.5 m. Data from all repetitions, including at 2 m and negative control, are available in the supplementary appendix.

**Figure 2.**
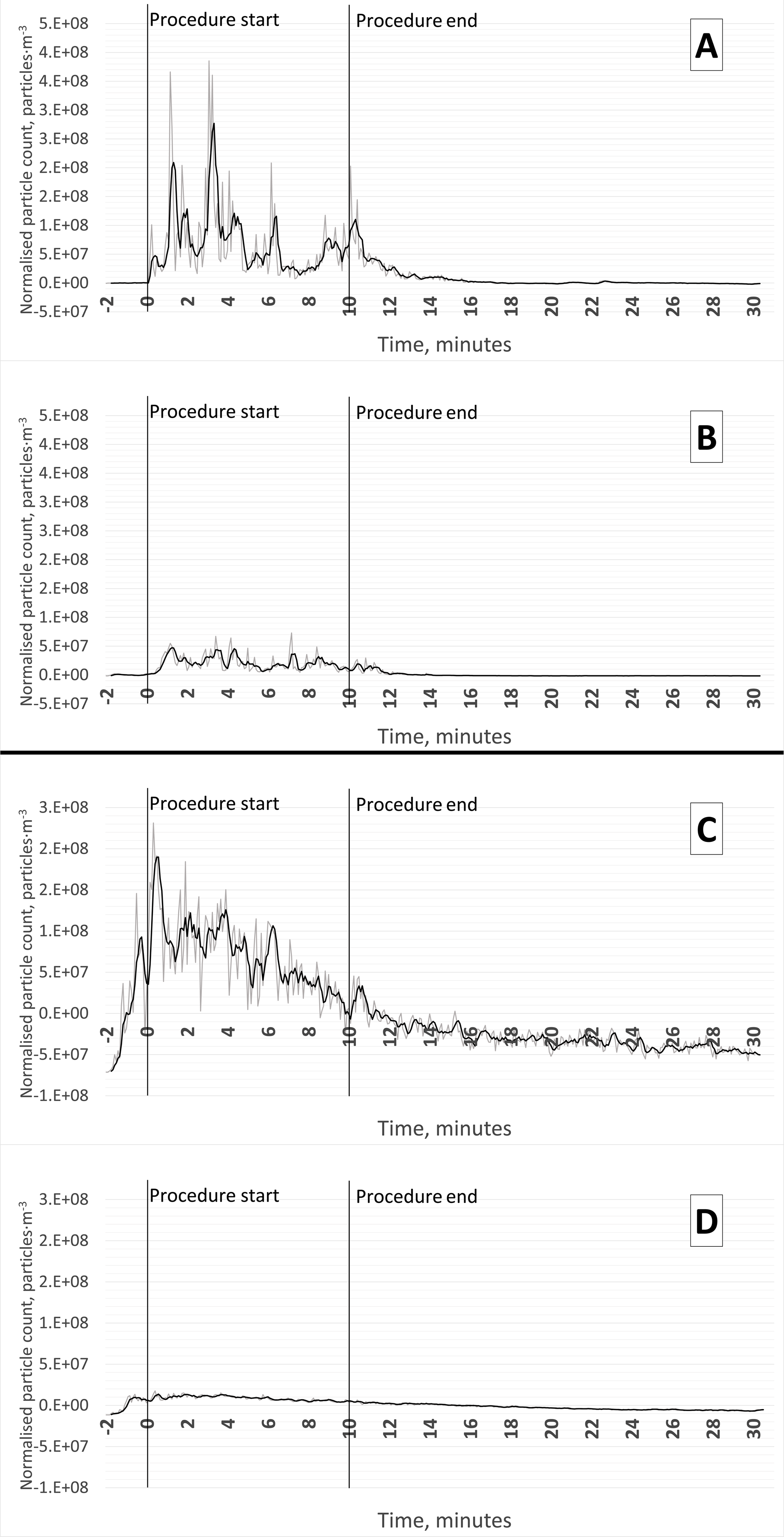
Suspended droplets measured using an optical particle counter at the 0.5 m location. Illustrative data are given in this figure from one repetition of each experiment, data from all repetitions are available in the supplementary appendix, as well as for 2 m and negative controls. The grey line represents the raw values, the black line represents a four-period moving average. A: Positive control with suction (no LEV) using the air-turbine handpiece in the open plan setting; B: Air-turbine with suction and LEV in the open plan setting; C: Positive control (no LEV or suction) using the ultrasonic scaler in the single surgery setting; D: Ultrasonic scaler with LEV only in the single surgery setting.

#### Active sampling with BioSamplers and fluorometric analysis

Detection of fluorescein decreased with increasing distance from the procedure. The use of LEV was associated with a 75 – 91% reduction in aerosolised fluorescein from the air-turbine handpiece dependant on location. The percentage reduction decreased with increasing distance from the dental procedure, and this was 90% within 0.5 m; this distance represents the breathing zone of members of the dental team (Table 1 and Figure 3).

**Table 1.**
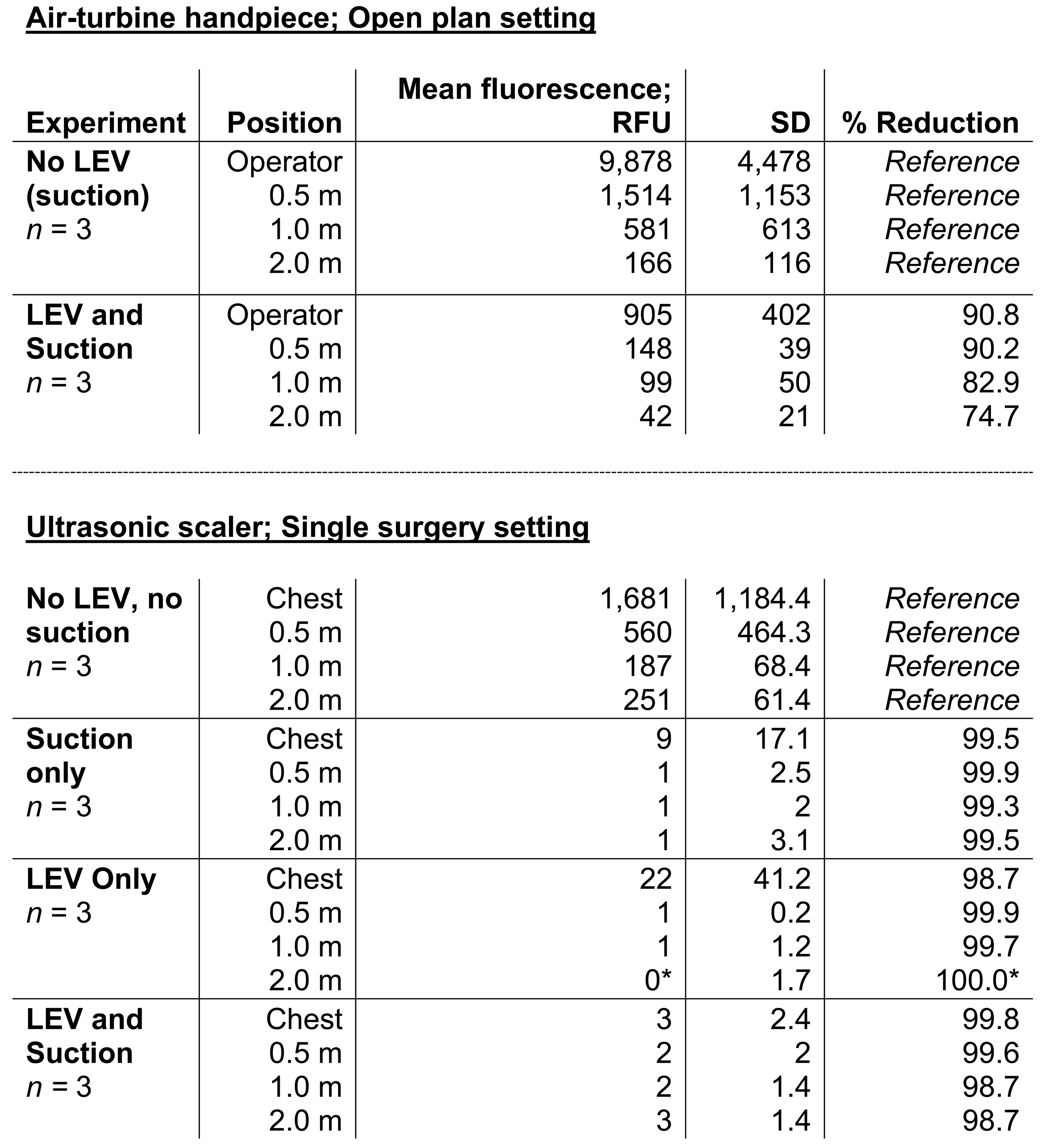
Aerosolised fluorescein collected by BioSampler and measured using fluorometric analysis. Data adjusted for background fluorescence by subtraction of the background reading from all data (open plan setting: 25.2 Relative Fluorescence Units, RFU, *n* = 12; single surgery setting: 25.8 RFU, *n* = 12). All air-turbine experiments also used dental suction. *Actual reading was below zero (−1 RFU) after subtraction of background reading but limited to zero for this table. LEV: Local Exhaust Ventilation.

Air-turbine handpiece; Open plan setting
| Experiment | Position | Mean fluorescence;<br>RFU | SD | % Reduction |
| --- | --- | --- | --- | --- |
| **No LEV**<br>**(suction)**<br>*n* = 3 | Operator | 9,878 | 4,478 | *Reference* |
|  | 0.5 m | 1,514 | 1,153 | *Reference* |
|  | 1.0 m | 581 | 613 | *Reference* |
|  | 2.0 m | 166 | 116 | *Reference* |
| **LEV and**<br>**Suction**<br>*n* = 3 | Operator | 905 | 402 | 90.8 |
|  | 0.5 m | 148 | 39 | 90.2 |
|  | 1.0 m | 99 | 50 | 82.9 |
|  | 2.0 m | 42 | 21 | 74.7 |

**Table 1.** Aerosolised fluorescein collected by BioSampler and measured using fluorometric analysis.
|  |  |  |  |  |
| --- | --- | --- | --- | --- |
| **No LEV, no**<br>**suction**<br>*n* = 3 | Chest | 1,681 | 1,184.4 | *Reference* |
|  | 0.5 m | 560 | 464.3 | *Reference* |
|  | 1.0 m | 187 | 68.4 | *Reference* |
|  | 2.0 m | 251 | 61.4 | *Reference* |
| **Suction**<br>**only**<br>*n* = 3 | Chest | 9 | 17.1 | 99.5 |
|  | 0.5 m | 1 | 2.5 | 99.9 |
|  | 1.0 m | 1 | 2 | 99.3 |
|  | 2.0 m | 1 | 3.1 | 99.5 |
| **LEV Only**<br>*n* = 3 | Chest | 22 | 41.2 | 98.7 |
|  | 0.5 m | 1 | 0.2 | 99.9 |
|  | 1.0 m | 1 | 1.2 | 99.7 |
|  | 2.0 m | 0* | 1.7 | 100.0* |
| **LEV and**<br>**Suction**<br>*n* = 3 | Chest | 3 | 2.4 | 99.8 |
|  | 0.5 m | 2 | 2 | 99.6 |
|  | 1.0 m | 2 | 1.4 | 98.7 |
|  | 2.0 m | 3 | 1.4 | 98.7 |

**Figure 3.**
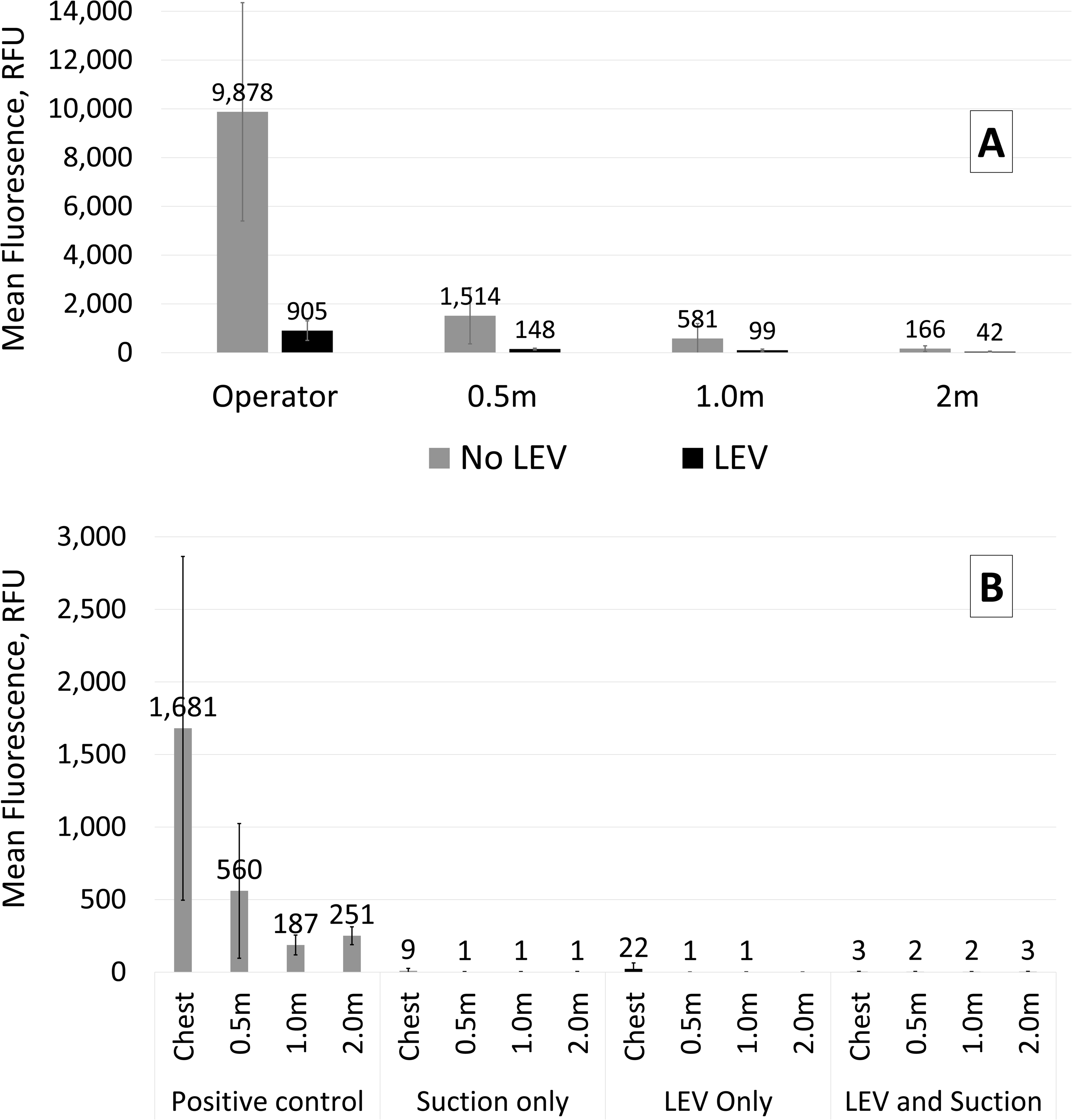
Aerosolised fluorescein collected by BioSampler and measured using fluorometric analysis. Error bars show 1SD in each direction. A: Experiments using the air-turbine handpiece in the open plan setting (all with suction). Data adjusted for background fluorescence by subtraction of the background reading (25.2 Relative Fluorescence Units, RFU; *n* = 12) from all data. B: Experiments using the ultrasonic scaler in the single surgery setting. Data adjusted for background fluorescence by subtraction of the background reading (25.8 RFU; *n* = 12) from all data.

#### Passive sampling

Samples were grouped by distance from the procedure: ≤ 0.5 m, 1 – 2 m, and 2.5 – 4 m. Sample RFU values were corrected for background fluorescence by subtracting the mean [SD] background RFU reading from each location (≤ 0.5m = 41 [20]; 1 – 2m = 41 [155]; 2.5 – 4m = 39 [21]) before calculating mean corrected RFU for each location. Values at 1 – 2 m and 2.5 – 4 m were substantially lower than at ≤ 0.5 m (Table 2 and Appendix Figure 9). At 0.5 m there was a 95% reduction in settled fluorescein when LEV was used. Between 1 – 2m there was a 69% reduction and at 2.5 – 4m this was 78%.

**Table 2.**
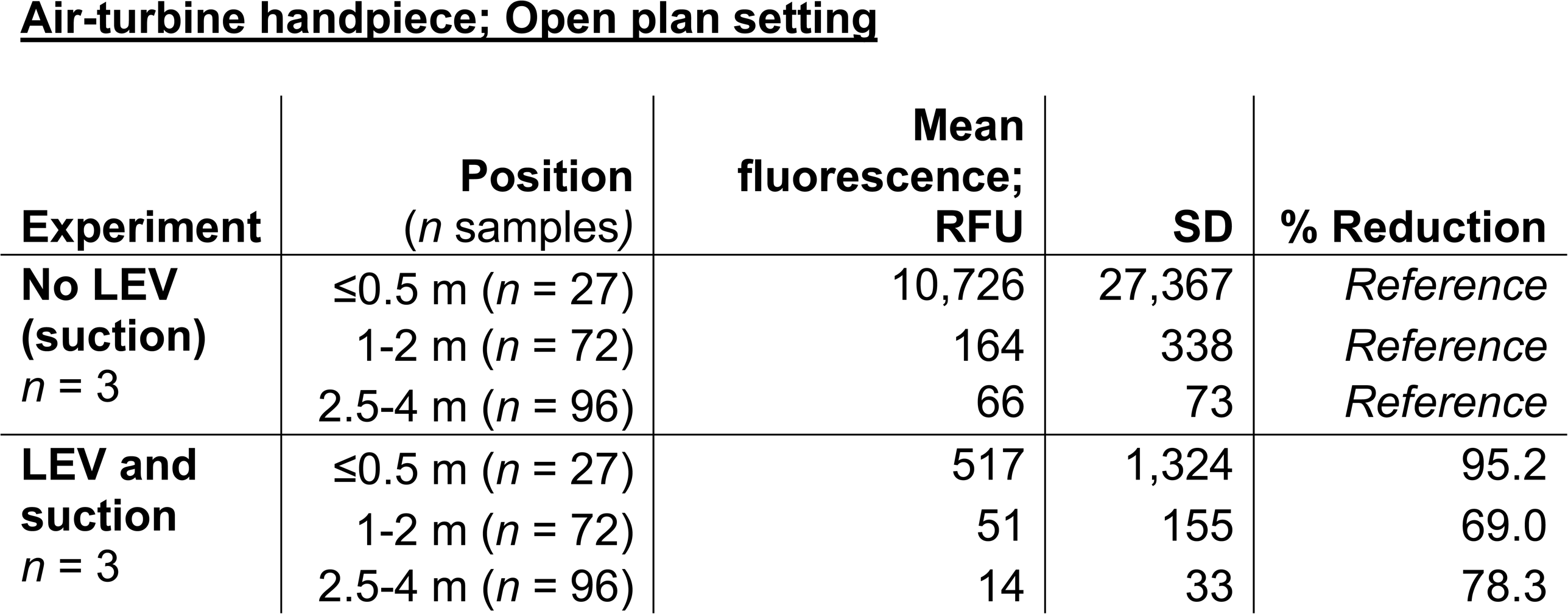
Fluorescein tracer from the air-turbine handpiece, collected by settlement onto filter paper samples in the open plan setting and measured using fluorometric analysis. Data for each group adjusted for background fluorescence in the respective location by subtraction of mean negative control values from each sample (≤ 0.5m = 41 RFU, 1-2m = 41 RFU, 2.5-4m = 39 RFU) before averaging. All experiments also used dental suction. RFU: Relative Fluorescence Units; LEV: Local Exhaust Ventilation.

| Experiment | Position<br>(*n* samples) | Mean<br>fluorescence;<br>RFU | SD | % Reduction |
| --- | --- | --- | --- | --- |
| **No LEV**<br>**(suction)**<br>*n* = 3 | ≤0.5 m (*n* = 27) | 10,726 | 27,367 | *Reference* |
|  | 1-2 m (*n* = 72) | 164 | 338 | *Reference* |
|  | 2.5-4 m (*n* = 96) | 66 | 73 | *Reference* |
| **LEV and**<br>**suction**<br>*n* = 3 | ≤0.5 m (*n* = 27) | 517 | 1,324 | 95.2 |
|  | 1-2 m (*n* = 72) | 51 | 155 | 69.0 |
|  | 2.5-4 m (*n* = 96) | 14 | 33 | 78.3 |

### Single surgery setting with ultrasonic scaler

The mean (SD; minimum – maximum) temperature during experiments was 24.1 °C (0.8; 22.7 – 26.4 °C) and relative humidity was 38.5 % (6.2; 26.0 – 45.0 %).

#### Active sampling with optical particle counters

Particle counts were substantially lower during all conditions at 2 m compared to 0.5 m. At 0.5 m and 2 m, the use of LEV was associated with a substantial reduction in particle counts. Figure 2 shows illustrative data from one repetition at 0.5 m. Data from all repetitions, including 2 m and negative control, are available in the supplementary appendix.

#### Active sampling with BioSamplers and fluorometric analysis

Detection of aerosolised fluorescein decreased with increasing distance from the procedure (Table 1 and Figure 3). At all locations, LEV was associated with a 98.7 – 100.0% reduction in aerosolised fluorescein.

## Discussion

Overall, three complementary sets of data at multiple locations, with different dental procedures across two clinical settings robustly demonstrate that LEV is effective in capturing aerosols and droplets from dental procedures and reducing dispersion. This reduction was most significant closest to the procedure, in the breathing zone of the operator and assistant.

Previous studies have evaluated the effectiveness of LEV in dentistry by studying droplet dispersion using a non-fluorescent tracer (Shahdad et al. 2020) and aerosols using particle counting instruments alone (Ehtezazi et al. 2021). These studies demonstrate substantial reductions in respective measures with LEV, however the present study is the first to examine the effect of LEV on both settled droplets and suspended aerosols simultaneously, and the first to capture suspended aerosols with a tracer specific to dental procedures (i.e., not potentially from another source as when measuring particles only). The positioning of LEV in the above studies was also more distant from the source (15 – 20 cm), whereas in the present study positioning was optimal for aerosol capture (10 cm). Relative reduction in aerosol was most pronounced for the ultrasonic scaler and we hypothesise that this is because the ultrasonic device produces particles with less momentum than those forced out under compressed air from the air-turbine; we propose that particles from the scaler are therefore more easily captured, explaining the more marked reduction.

We assessed the effectiveness of LEV for aerosol containment during dental procedures and we used dental suction during experiments to simulate standard practice. Previous studies demonstrate the significant benefit of dental suction (Allison et al. 2021a; Holliday et al. 2021) and the present study clearly demonstrates the *additional* benefit of LEV. In experiments with the air-turbine handpiece, dental suction was used during the control condition and with LEV, and the effect of LEV was marked even in addition to the effect of suction. With the ultrasonic scaler, the effect of suction was also measured separately to LEV, and here the effect of LEV with suction was similar to the effect of LEV alone; however, it was difficult to measure the effect of LEV in addition to suction due to how substantial the effect of suction was alone, with the scaler. This supports the hypothesis that particles from an ultrasonic scaler are more easily controlled with suction and LEV than those from an air-turbine. Importantly, the effect of dental suction may vary depending on the performance of each individual dental vacuum system and the actions of the operator; this is not the case for LEV. Additionally, pathogens are captured by the device’s HEPA filter, which is usually not the case for dental suction.

This study did not assess the practicality of using LEV for routine dentistry or the acceptability of the device for patients; however, in the authors’ opinion, the device is unobtrusive and there are unlikely to be significant barriers to use other than the floor space required and the need for decontamination. However, buying a device incurs initial costs and recurring costs for increased energy consumption, consumables and the safe disposal thereof. This is particularly relevant in multi-chair settings where several devices may be required.

The present study was conducted using a dental mannequin, and patients’ respiratory activities, which are significant aerosol sources (Wilson et al. 2021), were therefore not modelled. However, the study aimed to understand the effect of the LEV on the *additional* aerosols produced by the dental procedure, over and above normal clinical contact—an experimental design using a mannequin is ideal to allow this. The tracer showed where any aerosols from dental instruments were distributed to and the effect of LEV on these. Clearly, it is not the instrument aerosols themselves which pose a risk of infection, but the pathogens from *saliva* carried within these aerosols. Our previous work has shown that ‘saliva’, modelled with fluorescein tracer, is dispersed by aerosols from dental instruments (Holliday et al. 2021; Llandro et al. 2021). We chose to measure the aerosols from instruments themselves as dispersed ‘saliva’ is likely to be highly diluted; the model used in the present investigation therefore allowed us to demonstrate the effect of LEV with greater sensitivity than if a ‘saliva’-based model were used. The use of a fluorescent tracer is a reasonably straightforward approach to examine the distribution of dental aerosols, however the biological characteristics of *bioaerosols* cannot be examined, such as the infectivity of dispersed pathogens. Future studies should utilise biological tracers to validate findings from non-biological models such as those in the present study.

Particle counts from 0.3 – 10 μm OPC channels were combined, as this provides an easily comparable measure across experiments, and is consistent with measures used in air-quality monitoring combining particles < 10 μm, for example, PM_10_ (although this uses particle mass instead of number as in the present study). It is likely that particles of differing size behave differently; however, it was not the aim of the present study to examine this.

## Conclusion

This study demonstrates that LEV reduces aerosols from dental procedures by at least 90% within 0.5 m. Whilst no mitigation measure alone will completely eliminate risk, LEV appears to be a useful approach, which in addition to other measures, substantially reduces dispersion of aerosols, and therefore risk of exposure to pathogens. LEV seems more effective at capturing aerosols from ultrasonic scalers with less energetic droplets, compared to with an air-turbine handpiece, however, the effect remained substantial for the latter. LEV therefore shows promise in reducing aerosols from dental procedures and should play a role in reducing risks from dental bioaerosols.

## Data Availability

Full datasets are available at https://doi.org/10.25405/data.ncl.14987574

## Conflict of interest statement

This work was commissioned and funded by VODEX Ltd., Southampton, UK. The research was undertaken by independent investigators at the School of Dental Sciences, Newcastle University. VODEX Ltd. did not have any input into the design or conduct of this study, or the content of the manuscript. JRA, JD, NSJ, and RH have received related grants from the British Endodontic Society and the Royal College of Surgeons of Edinburgh which did not fund the present work. CD is funded by a National Institute of Health Research (NIHR) Newcastle Biomedical Research Centre Studentship. The views expressed are those of the authors and not necessarily those of VODEX Ltd., the National Health Service, NIHR or Department of Health and Social Care.

We would like to thank Philippa McClen for her support with this work. Contribution: JRA, RH—conception, design, data acquisition and interpretation, drafted and critically revised the manuscript; CD, KP, GC—data acquisition and interpretation, critically revised the manuscript; JD, NSJ—conception, design, drafted and critically revised the manuscript

## Supplementary Appendix

### Methods

#### Pilot testing

Preliminary testing was carried out to ensure fluorescein was captured by the Local Exhaust Ventilation (LEV) device and not redistributed into the environment which could lead to spurious results. Three filter papers were placed 11 cm away from the two exhaust vents on both sides of the LEV device, and one BioSampler was placed 22 cm away from each vent (total 6 filter papers and 2 BioSamplers). The LEV was switched on, and after four minutes, an air turbine handpiece was operated with fluorescein tracer as described in the main document, with the spray directed into the LEV nozzle. The handpiece was used for six minutes before stopping, and after 10 further minutes the LEV was turned off and samples were collected. A negative control condition with plain water in clean waterlines instead of fluorescein was also conducted and both conditions were conducted in triplicate. Samples were processed as described in the main document.

### Results

#### Pilot testing

No difference was seen in fluorescein detection between the negative control and when fluorescein tracer was used for pilot experiments, confirming that fluorescein does not pass through the LEV’s HEPA filter. These data are shown in Appendix Table 2 and Appendix Figure 2.

**Appendix Figure 1.**
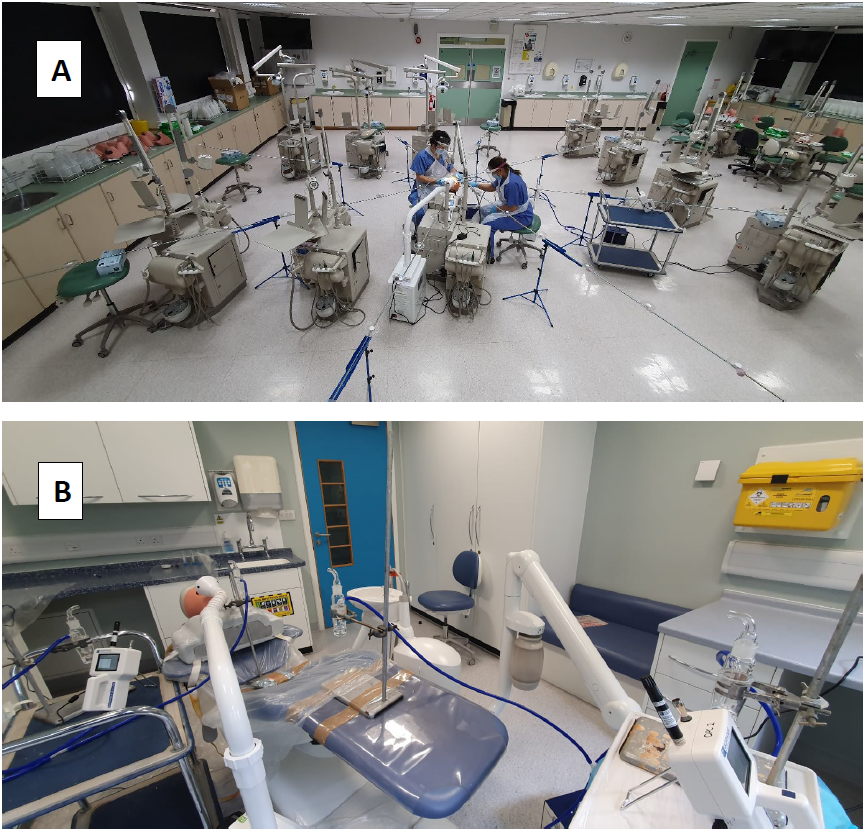
(A) Image of the experimental setup in the open plan setting showing the rig supporting filter papers. BioSamplers and Optical Particle Counter (OPC) number 1 are shown along the arm and trolley in the right of the image, OPC number 2 is adjacent to the LEV nozzle. (B) Image of the experimental setup in the single surgery setting showing BioSamplers placed along the axis of the dental chair, and OPCs placed at the foot of the dental chair and in the left of the image.

**Appendix Table 1.**
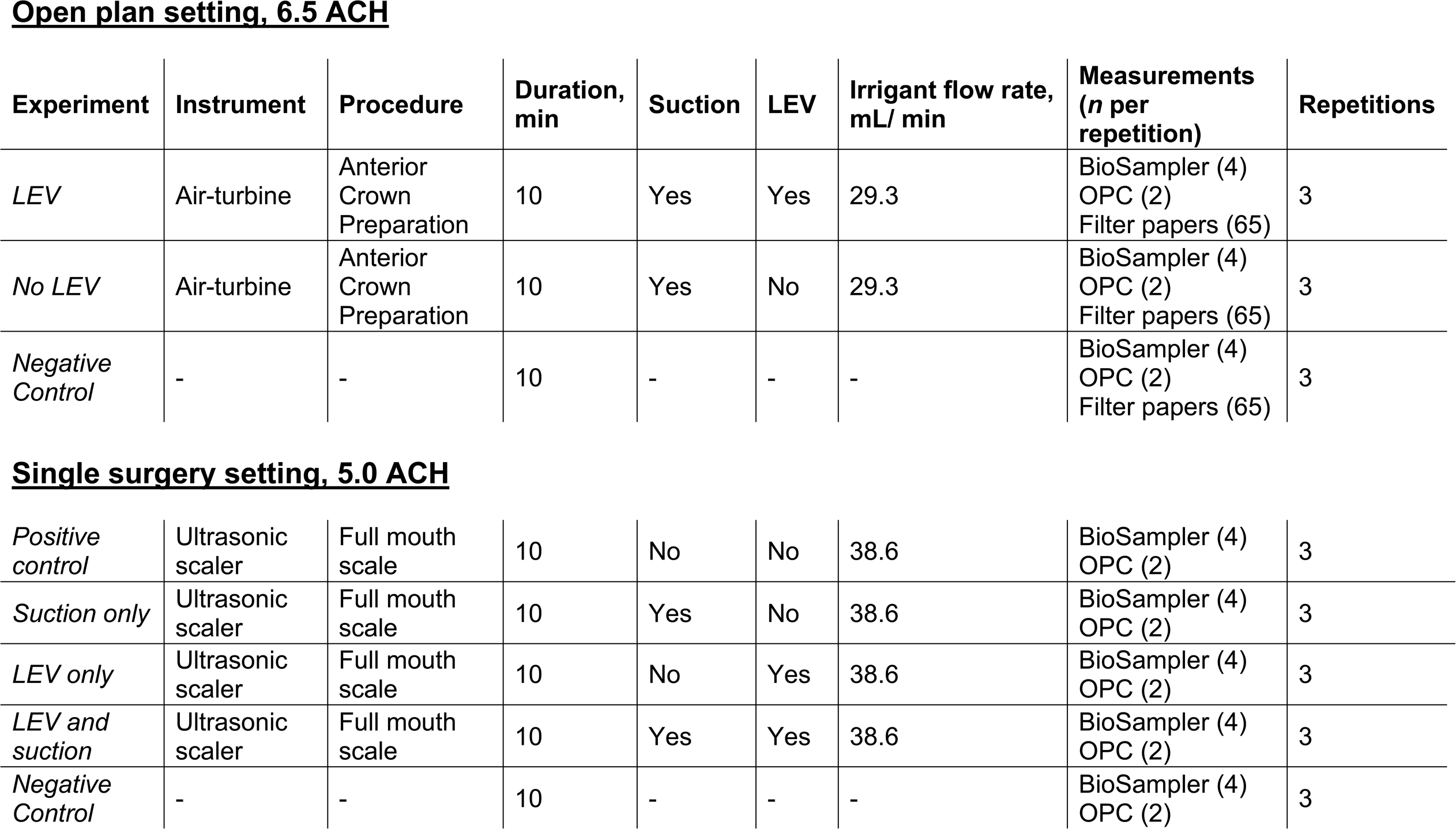
Overview of experiments conducted in each setting. ACH: Air Changes per Hour; LEV: Local Exhaust Ventilation; OPC: Optical Particle Counter.

**Open plan setting, 6.5 ACH**
| Experiment | Instrument | Procedure | Duration, min | Suction | LEV | Irrigant flow rate, mL/ min | Measurements (n per repetition) | Repetitions |
| --- | --- | --- | --- | --- | --- | --- | --- | --- |
| *LEV* | Air-turbine | Anterior Crown Preparation | 10 | Yes | Yes | 29.3 | BioSampler (4)<br>OPC (2)<br>Filter papers (65) | 3 |
| *No LEV* | Air-turbine | Anterior Crown Preparation | 10 | Yes | No | 29.3 | BioSampler (4)<br>OPC (2)<br>Filter papers (65) | 3 |
| *Negative Control* | - | - | 10 | - | - | - | BioSampler (4)<br>OPC (2)<br>Filter papers (65) | 3 |

|  |  |  |  |  |  |  |  |  |
| --- | --- | --- | --- | --- | --- | --- | --- | --- |
| *Positive control* | Ultrasonic scaler | Full mouth scale | 10 | No | No | 38.6 | BioSampler (4)<br>OPC (2) | 3 |
| *Suction only* | Ultrasonic scaler | Full mouth scale | 10 | Yes | No | 38.6 | BioSampler (4)<br>OPC (2) | 3 |
| *LEV only* | Ultrasonic scaler | Full mouth scale | 10 | No | Yes | 38.6 | BioSampler (4)<br>OPC (2) | 3 |
| *LEV and suction* | Ultrasonic scaler | Full mouth scale | 10 | Yes | Yes | 38.6 | BioSampler (4)<br>OPC (2) | 3 |
| *Negative Control* | - | - | 10 | - | - | - | BioSampler (4)<br>OPC (2) | 3 |

**Appendix Table 2.**
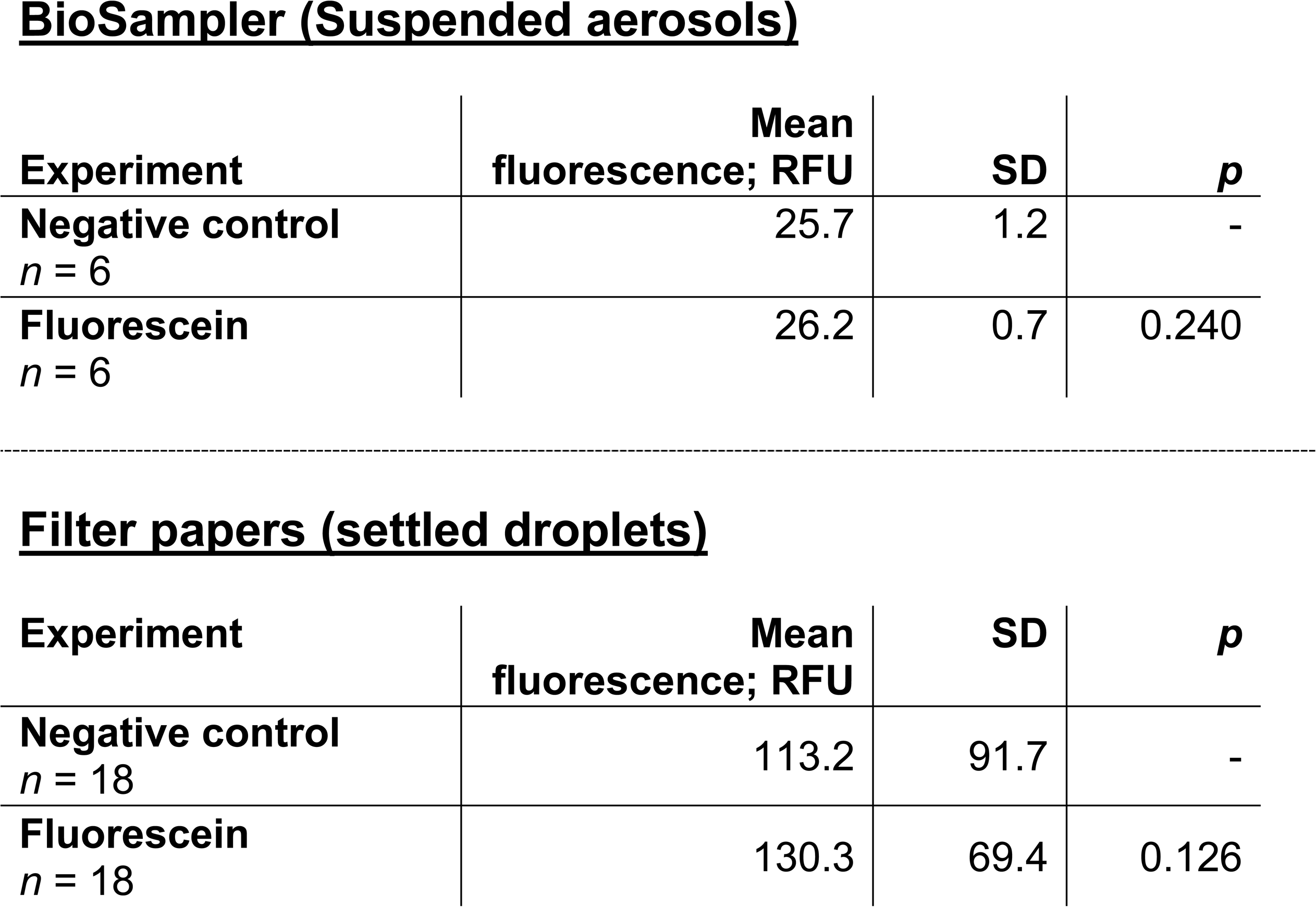
Fluorescence readings from pilot testing using filter papers and BioSamplers. The negative control condition used water instead of fluorescein. Data were non-normally distributed, and means were compared using the Man-Whitney U test with exact probabilities. RFU: Relative Fluorescence Units.

BioSampler (Suspended aerosols)
| Experiment | Mean<br>fluorescence; RFU | SD | *p* |
| --- | --- | --- | --- |
| Negative control<br>*n* = 6 | 25.7 | 1.2 | - |
| Fluorescein<br>*n* = 6 | 26.2 | 0.7 | 0.240 |

**Appendix Table 2.** Fluorescence readings from pilot testing using filter papers and BioSamplers.
| Experiment | Mean<br>fluorescence; RFU | SD | *p* |
| --- | --- | --- | --- |
| Negative control<br>*n* = 18 | 113.2 | 91.7 | - |
| Fluorescein<br>*n* = 18 | 130.3 | 69.4 | 0.126 |

**Appendix Figure 2.**
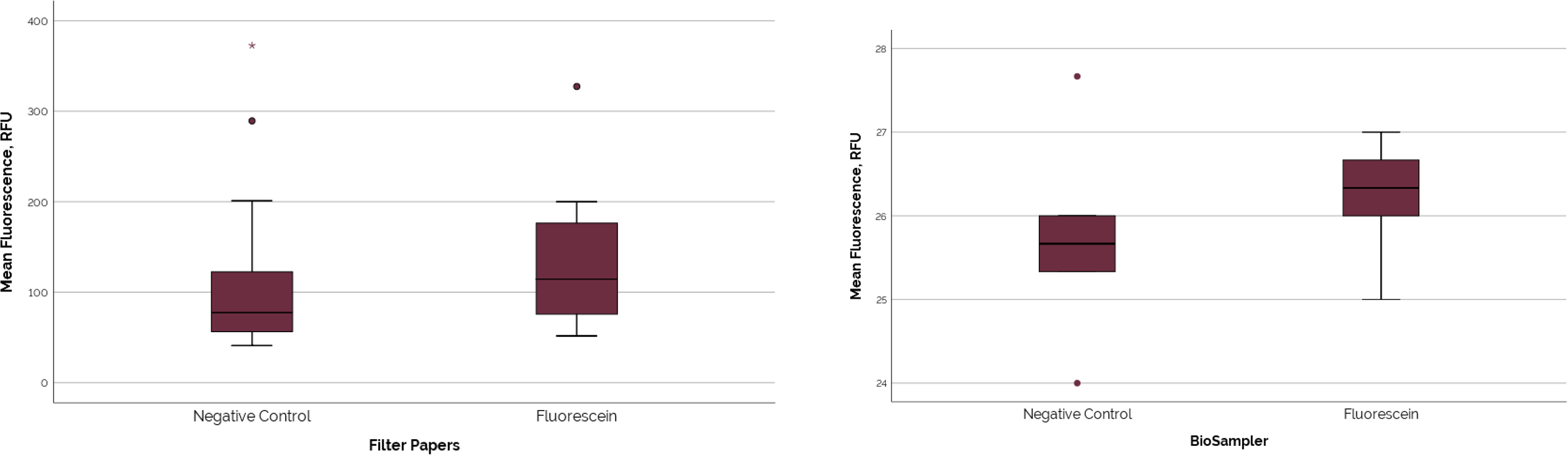
Box plots of fluorescence readings from pilot testing. Relative Fluorescence Units = RFU.

**Appendix Figure 3.**
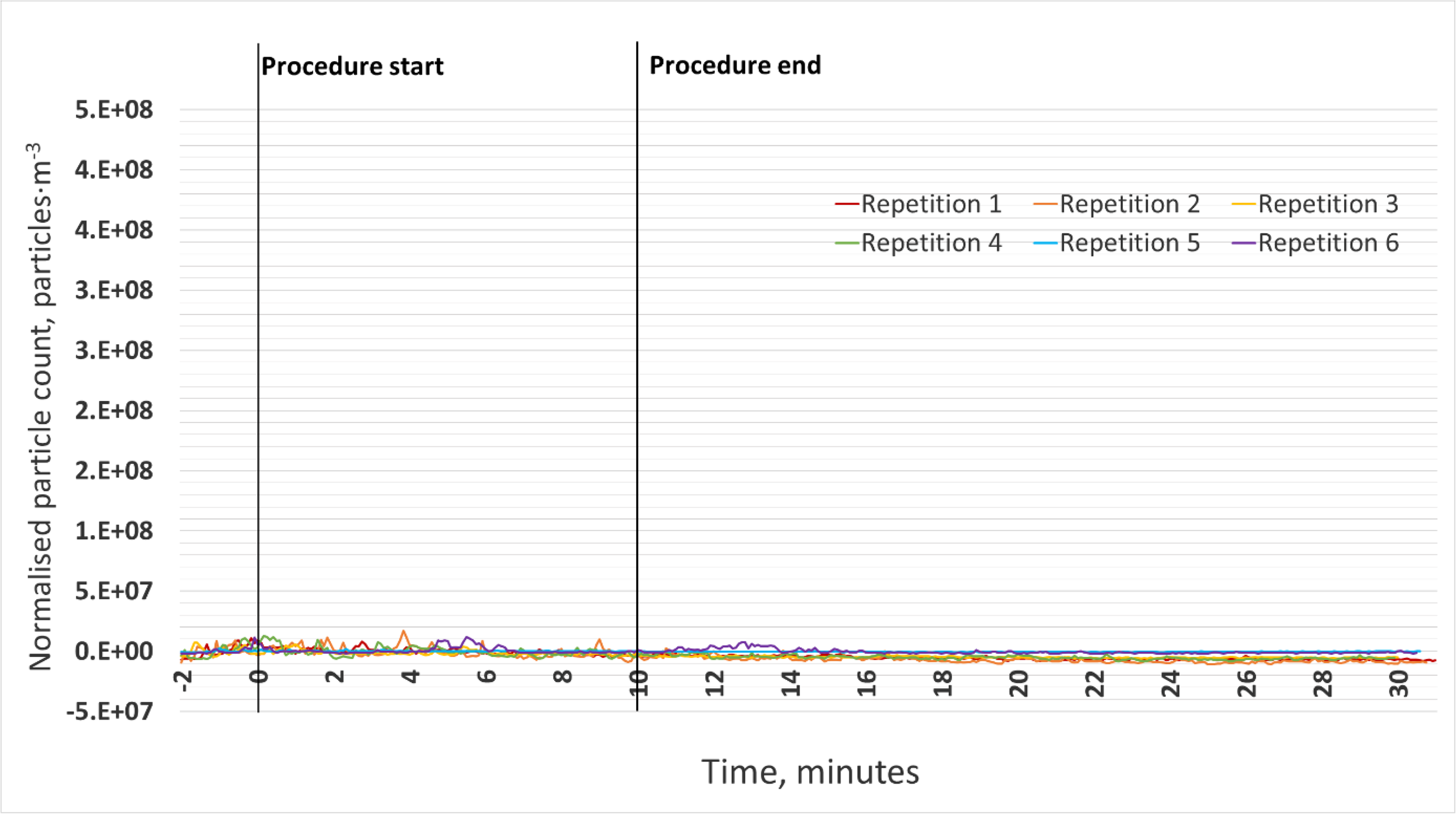
Negative control condition (*n* = 6) in the open plan setting where no dental procedure was taking place.

**Appendix Figure 4.**
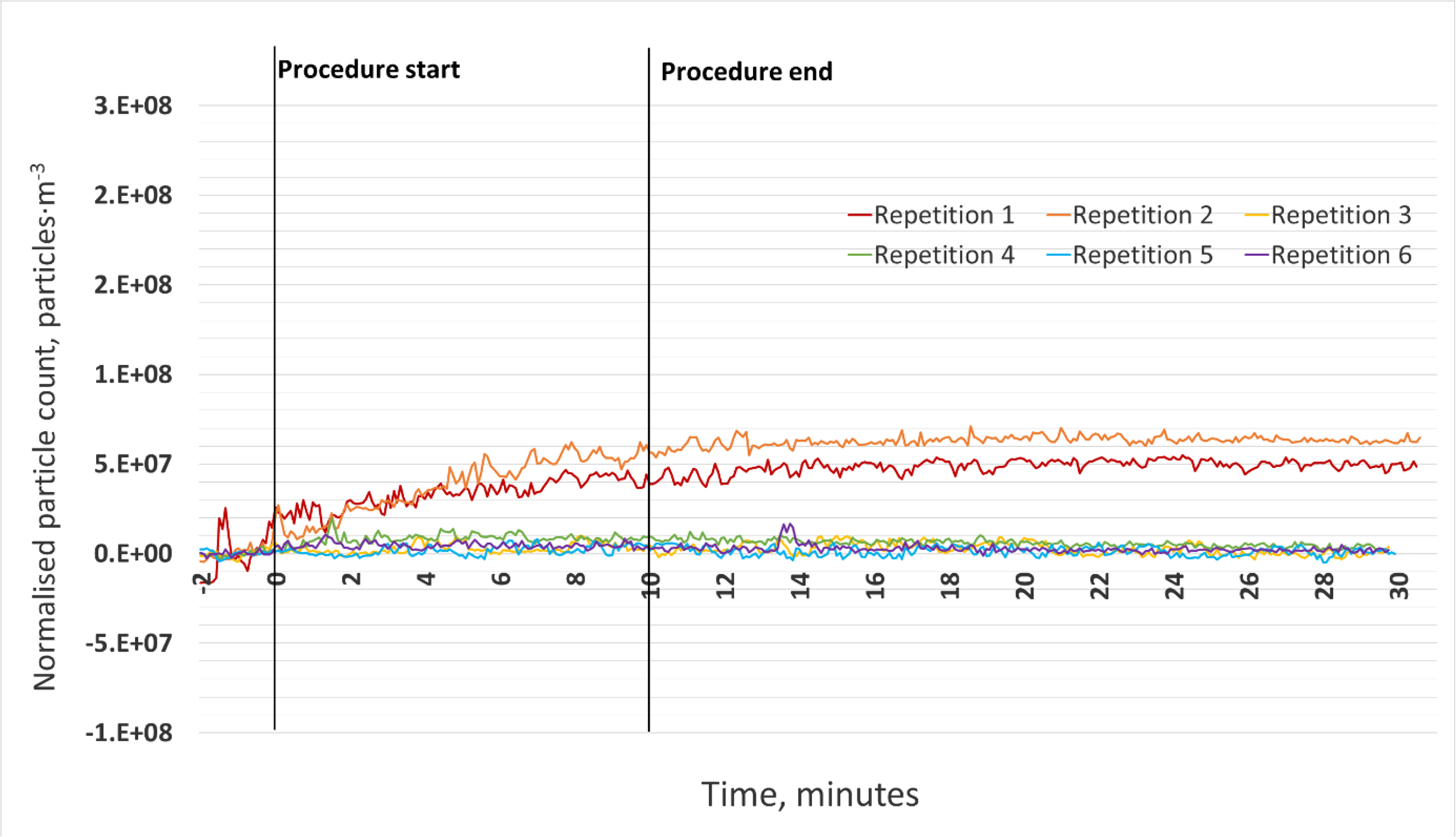
Negative control condition (*n* = 6) in the single surgery setting where no dental procedure was taking place. Raised counts in repetitions 1 and 2 likely represent signal drift as instruments equilibrate as these were the first runs.

**Appendix Figure 5.**
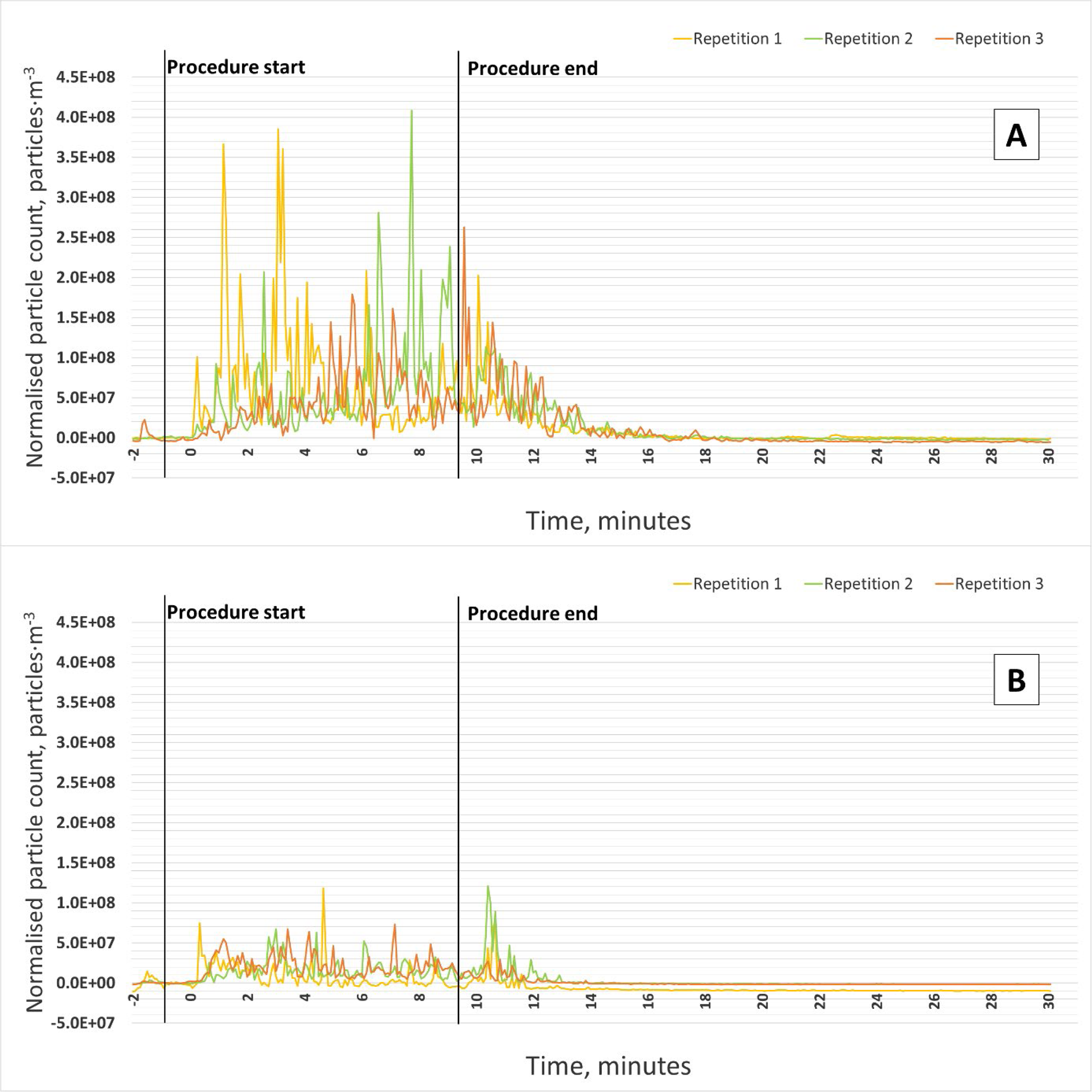
Suspended droplets from an air-turbine handpiece as measured by an optical particle counter. Data from three repetitions collected from the <u>0.5 m</u> sampling position in the open plan setting. A: Positive control (no LEV, with suction); B: LEV

**Appendix Figure 6.**
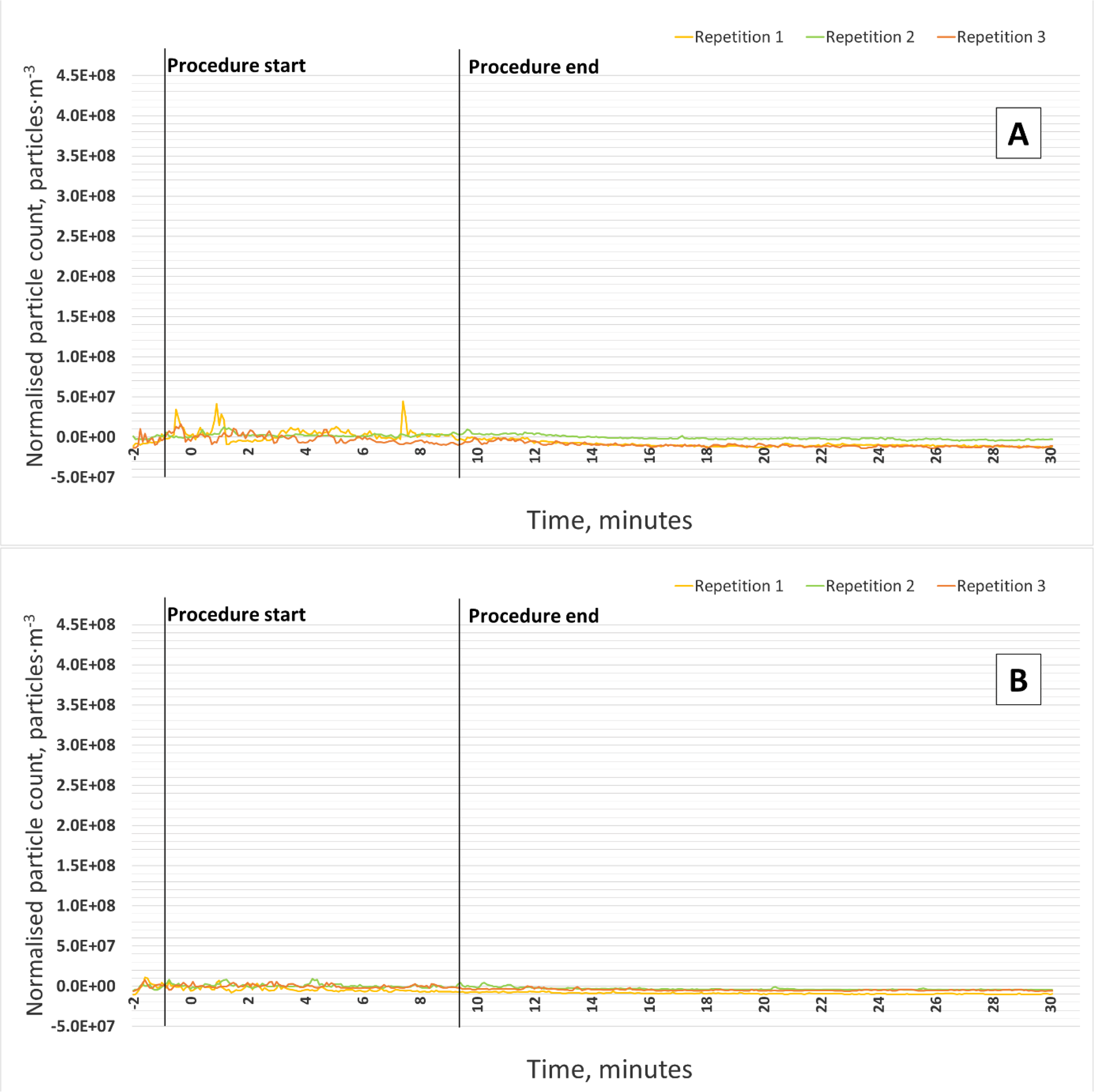
Suspended droplets from an air-turbine handpiece as measured by an optical particle counter. Data from three repetitions collected from the <u>2 m</u> sampling position in the open plan setting. A: Positive control (no LEV, with suction); B: LEV

**Appendix Figure 7.**
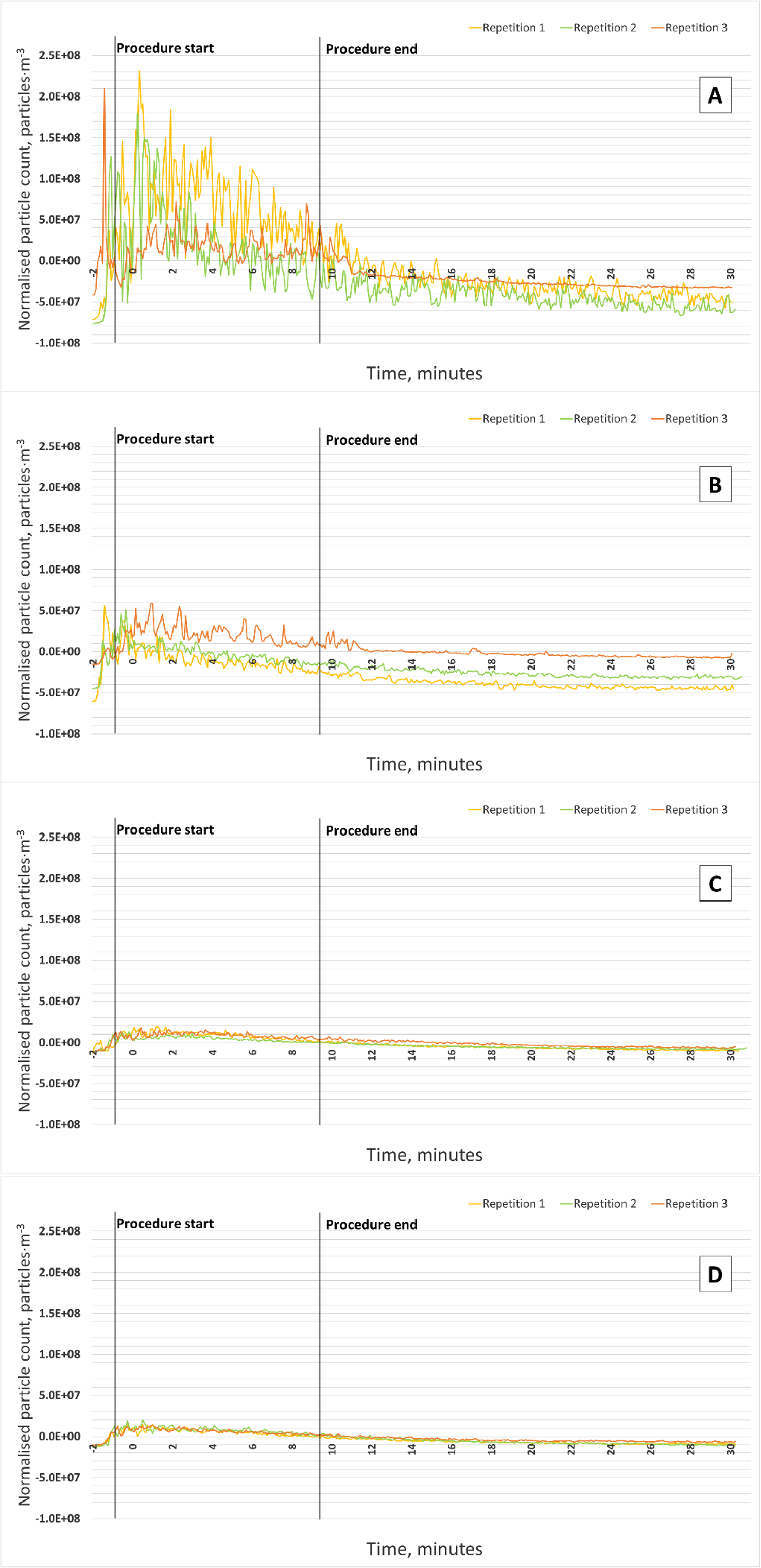
Suspended droplets from an ultrasonic scaler as measured by an optical particle counter. Data from three repetitions collected from the <u>0.5 m</u> sampling position in the single surgery setting. A: Positive control (no LEV or suction); B: Suction only; C: LEV only; D: LEV and suction.

**Appendix Figure 8.**
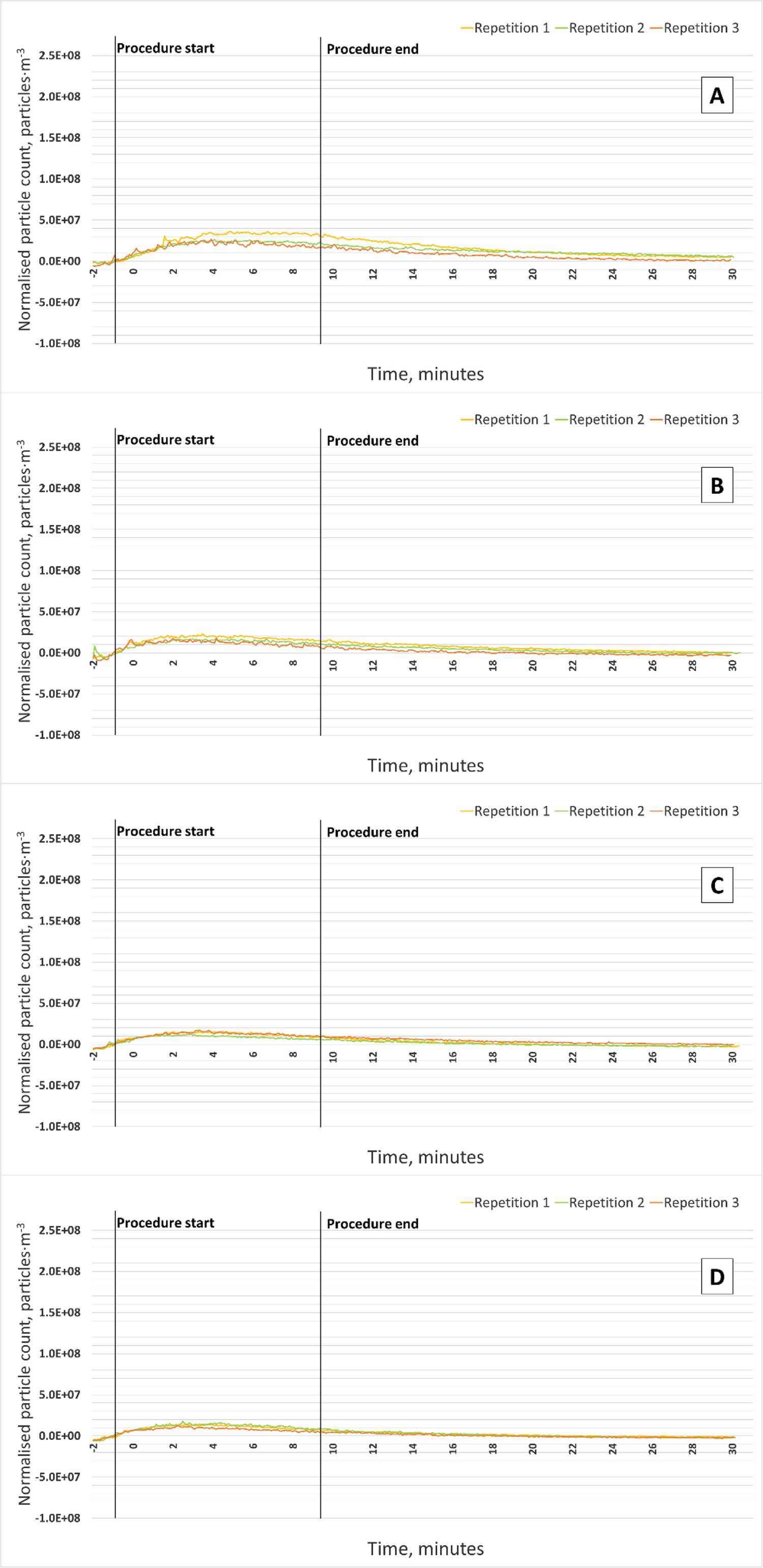
Suspended droplets from an ultrasonic scaler as measured by an optical particle counter. Data from three repetitions collected from the <u>2 m</u> sampling position in the single surgery setting. A: Positive control (no LEV or suction); B: Suction only; C: LEV only; D: LEV and suction.

**Appendix Figure 9.**
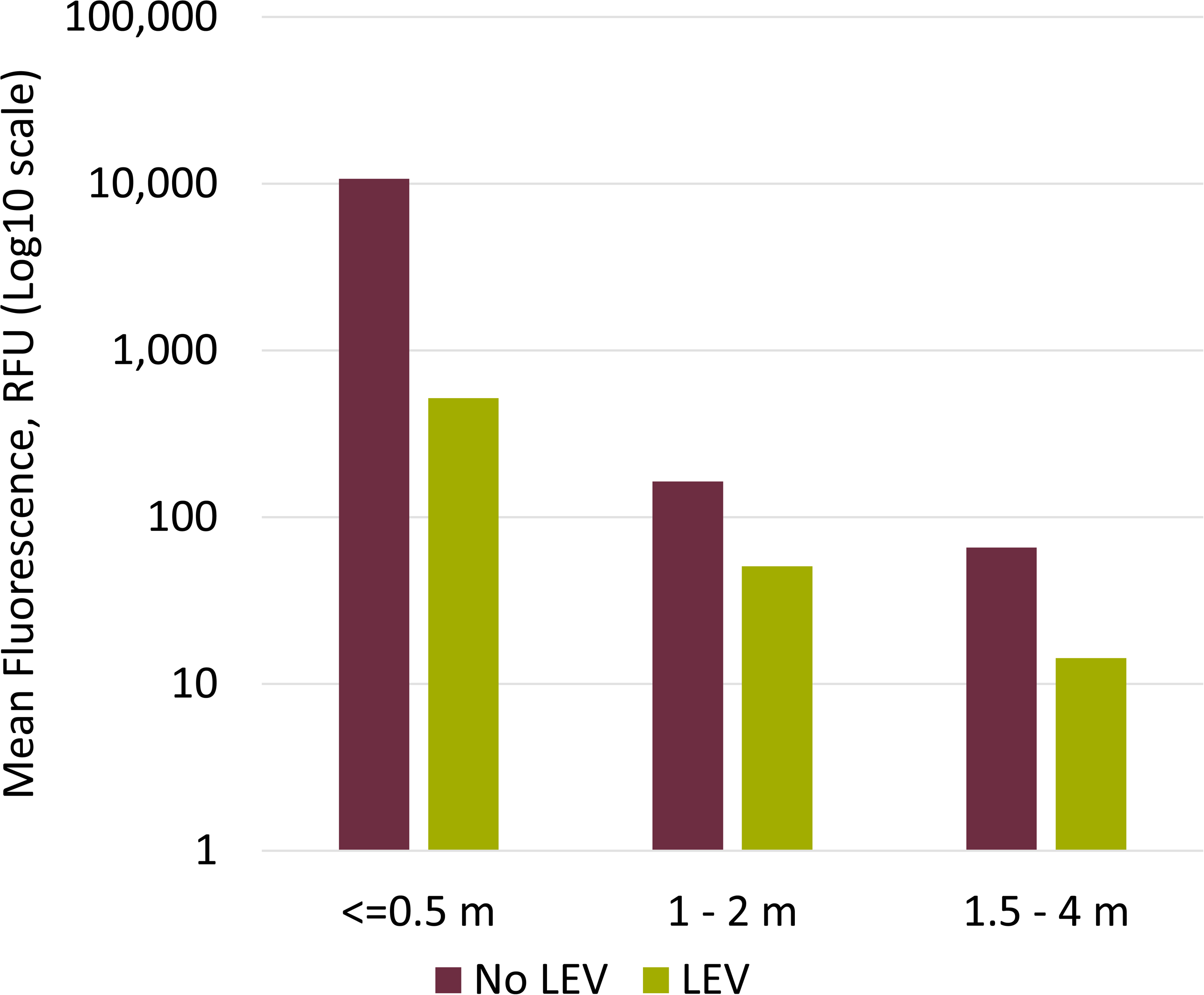
Fluorescein from the air-turbine handpiece, collected by settlement onto filter paper samples in the open plan setting and measured using fluorometric analysis. Data for each group adjusted for background fluorescence by subtraction of mean negative control values from each sample (≤ 0.5m = 41 RFU, 1-2m = 41 RFU, 2.5-4m = 39 RFU) before averaging. Error bars are not included as samples were pooled and large variation makes visualisation difficult. Standard deviations are given in Table 2 of the main manuscript. RFU: Relative Fluorescence Units.

